# Prognostic Nutritional Index in Risk of Mortality Following Fulminant Myocarditis

**DOI:** 10.1101/2024.07.22.24310842

**Authors:** Shunichi Doi, Yuki Ishibashi, Norio Suzuki, Daisuke Miyahara, Yukio Sato, Shingo Kuwata, Keisuke Kida, Masaki Izumo, Kenji Onoue, Koshiro Kanaoka, Yoshihiko Saito, Yoshihiro J. Akashi, Japanese Registry of Fulminant Myocarditis Investigators

## Abstract

**Background:** Fulminant myocarditis (FM) is an acute fatal inflammation disease, but its chronic phase is unclear. A Japanese nationwide registry evaluated the long-term mortality in FM patients using a prognostic nutritional index (PNI).

**Methods and Results:** The retrospective cohort study included patients with clinically suspected or histologically proven FM available for PNI. PNI was assessed on admission and at discharge. We divided patients into two groups based on PNI at discharge (PNI ≤40 or PNI >40) and analyzed the change in PNI and mortality between the groups. Of 323 patients (the median [first-third quartiles] age of this cohort was 50 [37–64] years, and 143 [44%] were female), PNI ≤40 at discharge was in 99 (31%) patients. The median PNI in all patients increased from 41 (36– 46) on admission to 43 (39–48) at discharge (*P*<0.0001). Patients with PNI ≤40 had a lower event-free rate of death or rehospitalization with cardiovascular causes than those with PNI >40 (log-rank *P*=0.0001). When the PNI at discharge, age, sex, left ventricular ejection fraction, and Barthel index were evaluated in a multivariable Cox regression analysis, PNI ≤40 had an independent association with the death or rehospitalization with cardiovascular causes (hazard ratio, 2.14 [95% confidence interval, 1.14–4.01]; *P*=0.0289).

**Conclusions:** One-third of FM patients with low PNI at discharge had a higher risk of mortality than those with high PNI in the chronic phase. This study provokes clinical insight into the phenotype of chronic inflammation in FM and optimal follow-up management with low PNI.

## Introduction

Fulminant myocarditis (FM) is a rare inflammation disease caused mainly by viruses, but it leads to a high mortality rate.^1,2^ Although mortality and readmission rates are lower in the chronic phase compared to the acute phase, several studies have suggested that the prognosis remains poor even in the chronic phase.^3,4,5^ FM patients with progression to cardiomyopathy in the chronic phase have been reported as the main indicator of prognosis.^6,7^ Inflammation of the myocardium can potentially persist, leading to severe fibrosis of the myocardium and chronic myocarditis. ^8,9,10^

The Prognostic Nutritional Index (PNI) not only suggests nutritional status but also indicates chronic inflammation^11^ and has been reported as a prognostic indicator for cancer, postoperative complications, and other inflammatory diseases.^12,13,14^ Recently, PNI has also been used to evaluate the prognosis of cardiovascular diseases, such as heart failure and myocardial infarction, because cardiovascular diseases may cause inflammation and have a worse prognosis.^15,16,17^

Few clinical studies have evaluated chronic inflammation in FM patients because FM is a rare and high-mortality disease in the acute phase. To our knowledge, no large-scale studies have evaluated the prognosis of FM using PNI. Therefore, we assessed the prognosis in the chronic phase of FM using PNI.

## Methods

### Study design and population

This study was a multicenter, retrospective, nationwide cohort study conducted in Japan. All the study participants were from the Japanese Registry of Fulminant Myocarditis (JRFM), which included patients with FM hospitalized in 235 facilities between April 2012 and March 2017, as previously described.^2^ We included patients aged ≥16 years who were diagnosed with FM based on the clinical diagnostic criteria of the European Society of Cardiology for clinically suspected myocarditis and the Japanese Circulation Society clinical diagnostic criteria for myocarditis.^18,19^ Patients with alternative diagnoses, such as ischemic heart disease, cardiomyopathy, and cardiac sarcoidosis, were not included in the cohort. To reveal the nutritional index in the post-acute phase, patients who died during hospitalization or had missing PNI data on admission and discharge were excluded. This study was conducted following the principles of the Declaration of Helsinki. The study protocol was approved by the Nara Medical University Ethics Committees (registration No.: 2256) in July 2019 and the Japanese Circulation Society (registration No.: 10) in November 2019. This study was registered at UMIN-CTR (University Hospital Medical Information Network Clinical Trials Registry registration No.: UMIN000039763).

### Nutritional status and outcomes

After we evaluated the patients’ discharge status, we clinically followed these patients and recorded their outcomes. To elucidate the characteristics and outcomes of patients with post-acute FM, we allocated patients with FM to 1 of 2 groups based on the PNI before discharge (PNI ≤40 and PNI >40 groups). The PNI cutoff was based on the lower cutoff value for the previous studies.^15,20^ We evaluated changes in PNI on admission and before discharge and outcomes according to the PNI category on discharge. PNI was calculated as 10×serum albumin (g/dL)+0.005×total lymphocyte count (per mm^3^).^21^ The primary endpoint was a time to first occurrence of the composite of all causes of death or nonelective hospitalizations for a primary cardiovascular reason, including heart failure, myocardial infarction, ventricular or atrial arrhythmias, and stroke. The total observation period was defined as the time between patients with discharge and loss of follow-up.

### Statistical analysis

Baseline clinical characteristics at hospital admission and discharge have been presented as numbers and percentages for categorical variables or median (first–third quartiles) for continuous variables. The Wilcoxon rank-sum test and Pearson χ2 test were used to compare continuous and categorical variables, respectively. We used the Wilcoxon matched-paired signed-rank test to analyze the paired PNI data at 2 different time points. We used the Kaplan-Meier method with log-rank tests and Cox regression models to analyze the outcomes for the total observational period. We assessed candidate patient characteristics from previous studies to determine prognostically relevant clinical characteristics.^2,7^ Cox regression analysis for prognostic factors was performed using multivariate imputation by chained equations based on 100 replications because some relevant characteristics were missing in this cohort. Using the Cox regression model, we estimated the HRs and 95% confidence intervals of outcomes. The follow-up period was determined by the last time point documented in the medical record, and patients who were lost to follow-up were regarded as censored without any events. All statistical tests were 2-sided, and P<0.05 was considered statistically significant. Statistical analyses were performed using the JMP Pro 16 software (SAS Institute Inc., Cary, NC, USA) and R version 4.3.2 for Windows.

## Results

### Overall study population and change of nutritional index

From the Japanese Registry of Fulminant Myocarditis cohort, we extracted data of 736 patients with histologically proven and clinically suspected myocarditis with fulminant presentation. In-hospital mortalities were 34% (247/736) in the patients. Of all patients who died during hospitalization, myocarditis was the direct cause of death for 210 (85%) patients. We included 323 patients (median age, 50 [37–64] years; female, 143 [44%]) who had available data to calculate the PNI on admission and at discharge (Figure 1). The median length of hospital stay was 29 (19– 49) days. Of 323 patients, 202 (63%) were with histologically proven FM.

**Figure 1.**
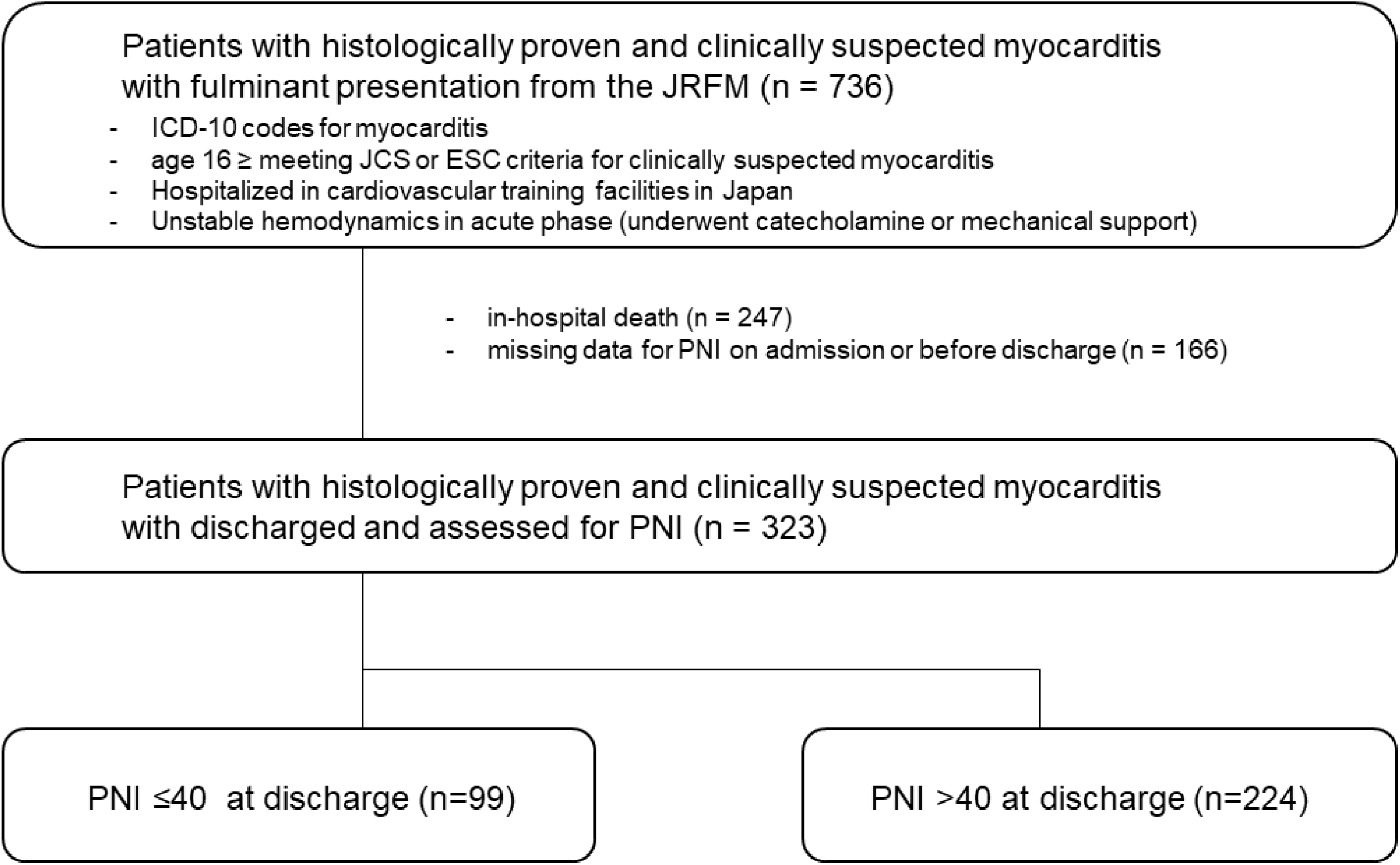
Flowchart over patient selection. JRFM, Japanese Registry of Fulminant Myocarditis; ICD-10, International Classification of Diseases–10; JCS, Japanese Circulation Society; ESC, indicates European Society of Cardiology; PNI, prognostic nutritional index.

The median PNI in all patients increased from 41 (36– 46) on admission to 43 (39–48) at discharge (P<0.0001; Figure 2A). The patients with PNI ≤40 on admission had lower PNI at discharge (P<0.0001; Figure 2B). Of 323 patients, 42 (13%) patients changed from PNI >40 on admission to PNI ≤40 at discharge, and 89 (28%) patients changed from PNI ≤40 on admission to PNI >40 at discharge (Figure 2C).

**Figure 2.**
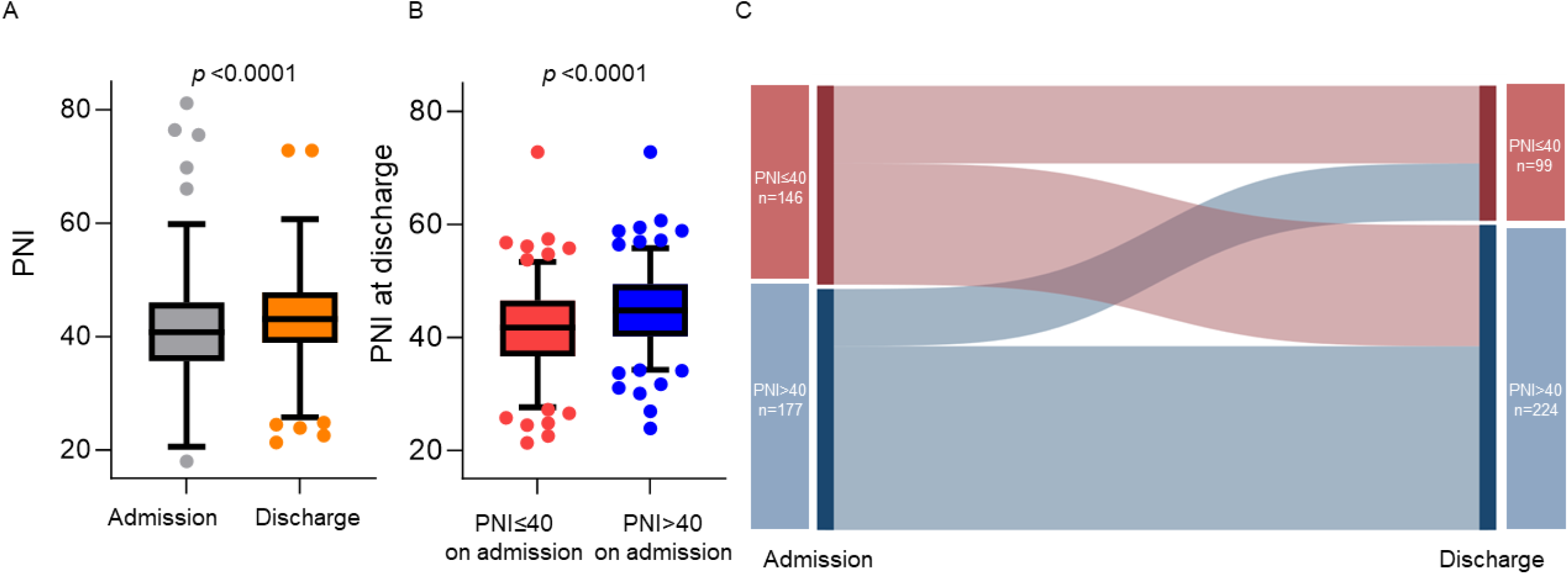
Comparison of the prognostic nutritional index on admission and at discharge. A, PNI increased at discharge from admission. B, Patients with PNI ≤40 on admission had lower PNI at discharge. C, Sankey diagram of PNI on admission and at discharge. PNI, prognostic nutritional index.

### Patient Characteristics and Outcomes According to PNI at Discharge

Of 323 patients, PNI >40 at discharge was in 224 (69%) patients, and PNI ≤40 in 99 (31%) patients (Table 1). The median ages were 45 (33–61) years and 60 (46–69) years, and women comprised 94 (42%) and 49 (49%) patients in the PNI >40 and PNI ≤40 groups, respectively. The patients with PNI ≤40 had a higher New York Heart Association (NYHA) classification, lower Barthel index on discharge, and a higher incidence of ventricular fibrillation. The lymphocytes, hemoglobin levels, albumin levels, estimated glomerular filtration rate (eGFR), and left ventricular ejection fraction (LVEF) were lower, and neutrocytes, C-reactive protein (CRP), and brain natriuretic peptide (BNP) were higher in PNI ≤40 groups compared with PNI >40 group. There was no significant difference between PNI >40 and PNI ≤40 groups in any medical history, treatment during hospitalization, temporary mechanical circulatory support devices, and length of hospital stay.

**Table 1.**
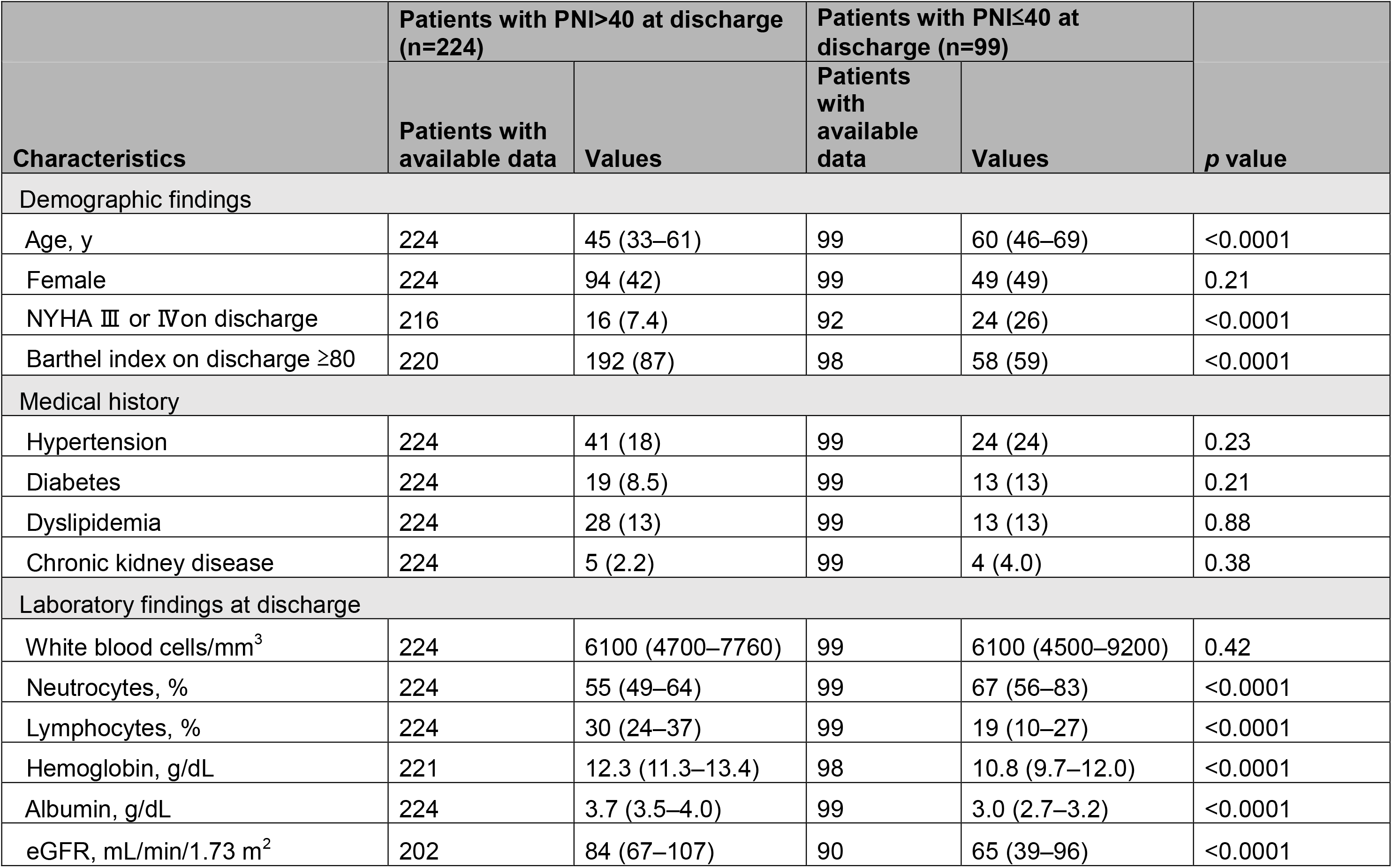
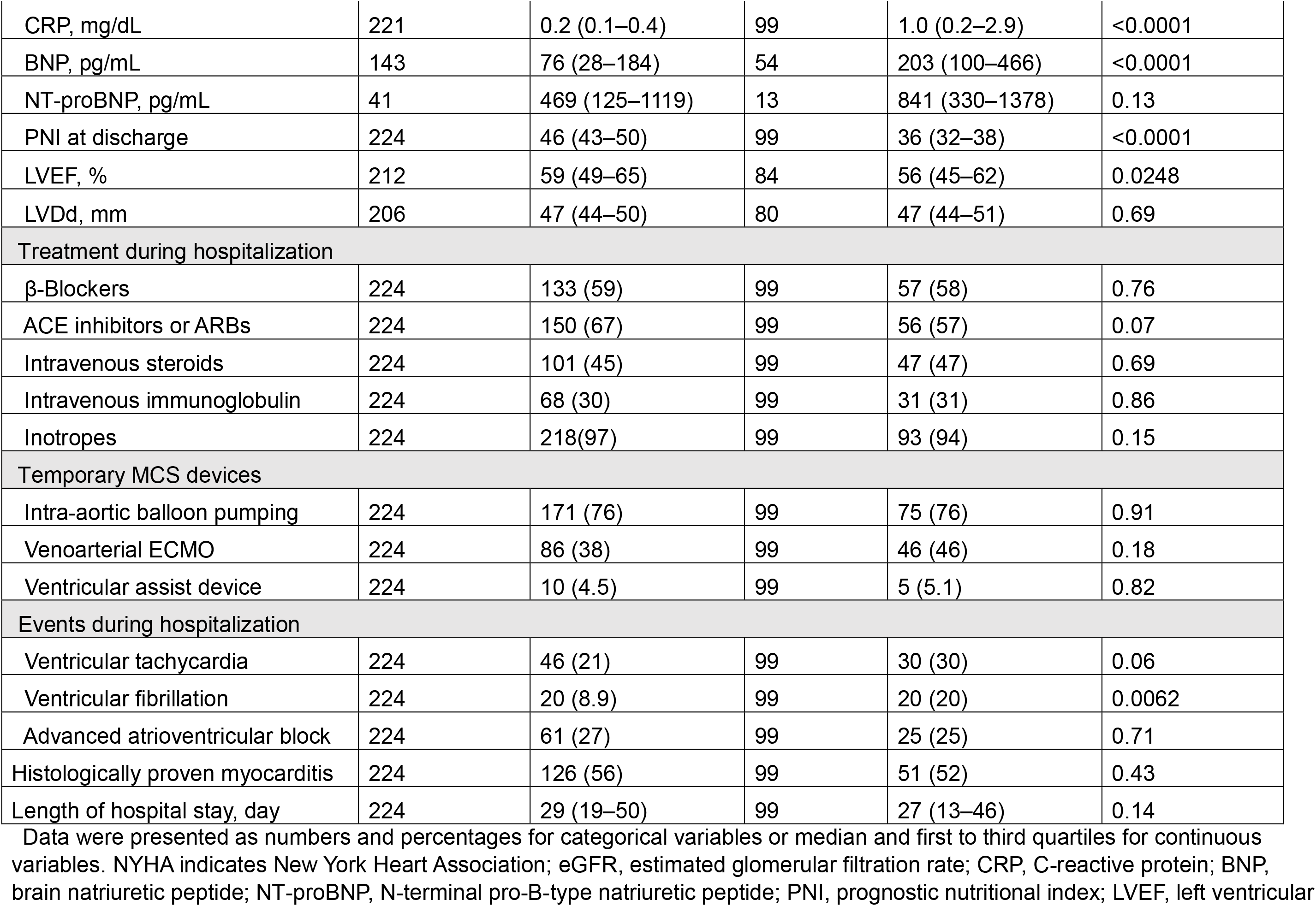
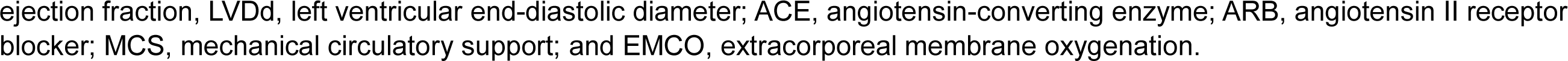
Characteristics of fulminant myocarditis at discharge.

| Characteristics | Patients with PNI>40 at discharge (n=224) |  | Patients with PNI≤40 at discharge (n=99) |  | p value |
| --- | --- | --- | --- | --- | --- |
|  | Patients with available data | Values | Patients with available data | Values |  |
| Demographic findings |  |  |  |  |  |
| Age, y | 224 | 45 (33–61) | 99 | 60 (46–69) | <0.0001 |
| Female | 224 | 94 (42) | 99 | 49 (49) | 0.21 |
| NYHA III or IV on discharge | 216 | 16 (7.4) | 92 | 24 (26) | <0.0001 |
| Barthel index on discharge ≥80 | 220 | 192 (87) | 98 | 58 (59) | <0.0001 |
| Medical history |  |  |  |  |  |
| Hypertension | 224 | 41 (18) | 99 | 24 (24) | 0.23 |
| Diabetes | 224 | 19 (8.5) | 99 | 13 (13) | 0.21 |
| Dyslipidemia | 224 | 28 (13) | 99 | 13 (13) | 0.88 |
| Chronic kidney disease | 224 | 5 (2.2) | 99 | 4 (4.0) | 0.38 |
| Laboratory findings at discharge |  |  |  |  |  |
| White blood cells/mm<sup>3</sup> | 224 | 6100 (4700–7760) | 99 | 6100 (4500–9200) | 0.42 |
| Neutrocytes, % | 224 | 55 (49–64) | 99 | 67 (56–83) | <0.0001 |
| Lymphocytes, % | 224 | 30 (24–37) | 99 | 19 (10–27) | <0.0001 |
| Hemoglobin, g/dL | 221 | 12.3 (11.3–13.4) | 98 | 10.8 (9.7–12.0) | <0.0001 |
| Albumin, g/dL | 224 | 3.7 (3.5–4.0) | 99 | 3.0 (2.7–3.2) | <0.0001 |
| eGFR, mL/min/1.73 m<sup>2</sup> | 202 | 84 (67–107) | 90 | 65 (39–96) | <0.0001 |
| CRP, mg/dL | 221 | 0.2 (0.1–0.4) | 99 | 1.0 (0.2–2.9) | <0.0001 |
| BNP, pg/mL | 143 | 76 (28–184) | 54 | 203 (100–466) | <0.0001 |
| NT-proBNP, pg/mL | 41 | 469 (125–1119) | 13 | 841 (330–1378) | 0.13 |
| PNI at discharge | 224 | 46 (43–50) | 99 | 36 (32–38) | <0.0001 |
| LVEF, % | 212 | 59 (49–65) | 84 | 56 (45–62) | 0.0248 |
| LVDd, mm | 206 | 47 (44–50) | 80 | 47 (44–51) | 0.69 |
| Treatment during hospitalization |  |  |  |  |  |
| β-Blockers | 224 | 133 (59) | 99 | 57 (58) | 0.76 |
| ACE inhibitors or ARBs | 224 | 150 (67) | 99 | 56 (57) | 0.07 |
| Intravenous steroids | 224 | 101 (45) | 99 | 47 (47) | 0.69 |
| Intravenous immunoglobulin | 224 | 68 (30) | 99 | 31 (31) | 0.86 |
| Inotropes | 224 | 218(97) | 99 | 93 (94) | 0.15 |
| Temporary MCS devices |  |  |  |  |  |
| Intra-aortic balloon pumping | 224 | 171 (76) | 99 | 75 (76) | 0.91 |
| Venoarterial ECMO | 224 | 86 (38) | 99 | 46 (46) | 0.18 |
| Ventricular assist device | 224 | 10 (4.5) | 99 | 5 (5.1) | 0.82 |
| Events during hospitalization |  |  |  |  |  |
| Ventricular tachycardia | 224 | 46 (21) | 99 | 30 (30) | 0.06 |
| Ventricular fibrillation | 224 | 20 (8.9) | 99 | 20 (20) | 0.0062 |
| Advanced atrioventricular block | 224 | 61 (27) | 99 | 25 (25) | 0.71 |
| Histologically proven myocarditis | 224 | 126 (56) | 99 | 51 (52) | 0.43 |
| Length of hospital stay, day | 224 | 29 (19–50) | 99 | 27 (13–46) | 0.14 |

Data were presented as numbers and percentages for categorical variables or median and first to third quartiles for continuous variables. NYHA indicates New York Heart Association; eGFR, estimated glomerular filtration rate; CRP, C-reactive protein; BNP, brain natriuretic peptide; NT-proBNP, N-terminal pro-B-type natriuretic peptide; PNI, prognostic nutritional index; LVEF, left ventricular
ejection fraction, LVDd, left ventricular end-diastolic diameter; ACE, angiotensin-converting enzyme; ARB, angiotensin II receptor blocker; MCS, mechanical circulatory support; and EMCO, extracorporeal membrane oxygenation.

The long-term outcomes are shown in Figure 3. The median follow-up period was 1114 (167–1667) days for all patients. The event-free rate of death or rehospitalization with cardiovascular causes at 1 and 3 years of follow-up was 94% and 90% in the PNI >40 groups and 79% and 73% in the PNI ≤40 group, respectively (log-rank P=0.0001; Figure 3A). During the 4-year observation, 21 patients died, and 22 patients were rehospitalized with cardiovascular causes. The PNI ≤40 group had a higher risk of mortality than the PNI >40 group (log-rank P<0.0001; Figure 3B), whereas there was no significant difference in rehospitalization with cardiovascular causes (log-rank P=0.72; Figure 3C). When the PNI, age, sex, LVEF, and Barthel index were evaluated in a multivariable Cox regression analysis, the PNI ≤40 showed an independent association with the primary endpoint(hazard ratio 2.14 [1.14–4.01], Table 2).

**Table 2.** Factors associated with all-cause death or rehospitalization with cardiovascular causes in fulminant myocarditis.

| Characteristics | Univariable |  | Multivariable |  |
| --- | --- | --- | --- | --- |
|  | HR (95% CI) | *P* value | HR (95% CI) | *P* value |
| Age, y, per 10 years | 1.23 (1.03–1.46) | 0.0210 | 1.20 (0.99–1.45) | 0.06 |
| Female | 0.97 (0.53–1.76) | 0.91 | 1.01 (0.54–1.89) | 0.97 |
| LVEF <50% at discharge | 3.78 (1.95–7.31) | 0.0002 | 3.70 (1.94–7.07) | 0.0002 |
| Barthel index ≥80 at discharge | 0.41 (0.22–0.78) | 0.0074 | 0.45 (0.23–0.88) | 0.0209 |
| PNI ≤40 at discharge | 3.00 (1.65–5.45) | 0.0006 | 2.14 (1.14–4.01) | 0.0191 |

LVEF indicates left ventricular ejection fraction; PNI, prognostic nutritional index; HR, hazard ratio; and CI, confidence interval

**Figure 3.**
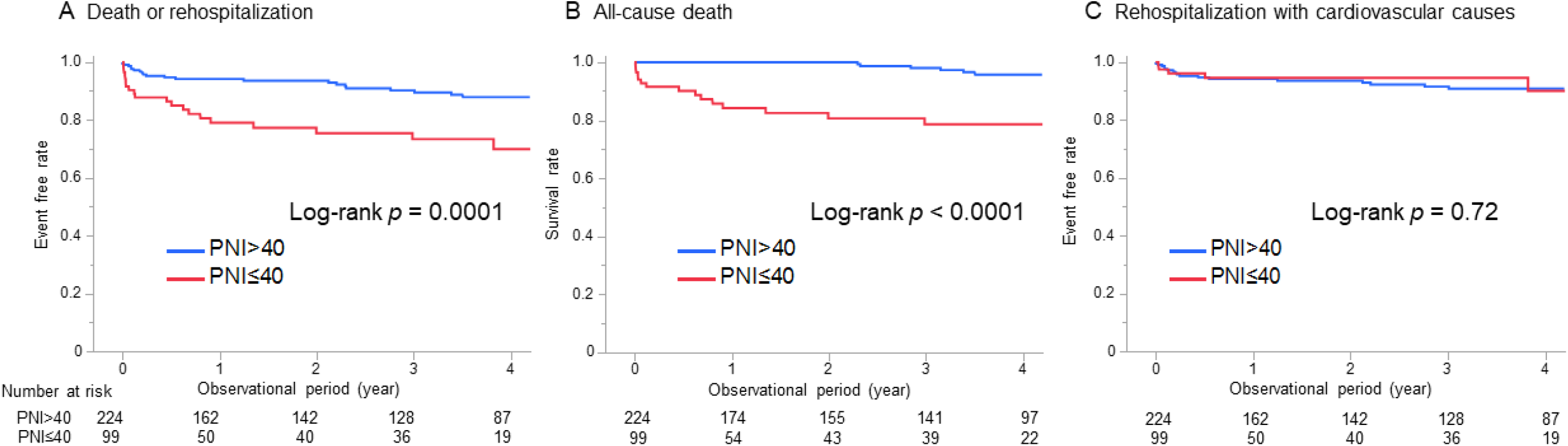
Kaplan-Meire curves for the outcomes. A, Event free rate of all-cause death or rehospitalization with cardiovascular causes between PNI ≤40 and PNI >40 at discharge. B, Survival rate between PNI ≤40 and PNI >40 at discharge. C, Event free rate of rehospitalization with cardiovascular causes between PNI ≤40 and PNI >40 at discharge. PNI, prognostic nutritional index.

### Patient Characteristics and Outcomes According to PNI on admission

Of 323 patients, PNI >40 on admission was in 177 (55%) patients, and PNI ≤40 on admission in 146 (45%) patients (Table S1). Patients with PNI ≤40 on admission were older, likely to be women, had higher NYHA class and had a prevalence of chronic kidney disease. The lymphocytes, hemoglobin levels, albumin levels, and eGFR were lower, and neutrocytes, CRP, and BNP were higher in patients with PNI ≤40 on admission than in patients with PNI >40 on admission. The patients with PNI ≤40 on admission had a longer hospital stay. In contrast, there was no significant difference between the groups with PNI >40 and PNI ≤40 on admission in any incidence of events, treatment during hospitalization, and temporary mechanical circulatory support devices. The delta values of albumin and lymphocytes between discharge and admission were significantly decreased in the PNI ≤40 group, respectively (P<0.0001, Figure S1). Patients with PNI ≤40 on admission had no significant incidence rate of death or rehospitalization with cardiovascular causes compared to those with PNI >40 on admission (log-rank P=0.26; Figure S2).

### Subgroup patients with histologically proven FM patients in PNI

Of 323 patients, 177 (55%) were histologically proven by endomyocardial biopsy (EMB). There was no significant difference, whether histologically proven FM, between PNI groups on admission or discharge (Table 1 and Table S1). PNI according to histological subtype are shown in Figure S3A. PNI was no significant difference between the histological subtype (p=0.15). The histologic severity of the acute phase of FM patients was evaluated centrally for this study using EMB specimens of 159 patients. EMB specimens had 109 (69%) degeneration and 77 (43%) necrosis. There was no significant difference in the PNI between histologic severity (Figure S3B and S3C).

## Discussion

In this largest nationwide multicenter cohort, discharged FM patients with low PNI had a higher risk of mortality. One-third of patients in clinical FM patients after discharge had low PNI, which indicates malnutrition, residual inflammation, and a worsening prognosis. PNI was an independent predictor of death or rehospitalization with cardiovascular causes in clinical FM patients with the chronic phase. Although low PNI on admission was associated with low PNI at discharge, PNI on admission could not be associated with mortality risk. The PNI, including albumin and lymphocytes, increased between admission and discharge. Although most myocarditis was assumed to be healed with inflammation, there is limited information on the long-term follow-up of residual inflammation in patients with FM after discharge. The first paper from the Japanese Registry of Fulminant Myocarditis reported the clinical findings based on the patient characteristics on admission,^2^ and we revealed the long-term mortality with PNI in clinical FM patients after discharge. This study provokes valuable clinical insight into the phenotype of chronic inflammation in FM and optimal follow-up management in FM patients with low PNI who are at a higher risk of mortality than patients with high PNI.

Myocarditis is induced predominantly by viruses or a wide variety of toxic substances and drugs.^22,23,24^ Although the aetiopathogenesis, induction, and course of myocarditis related to different infectious agents vary considerably, all myocarditis have inflammation and could injure the cardiac function and structure. Recently, three types of clinical possibilities with inflammation of viral myocarditis in the chronic phase have been recognized: (1) the virus is completely cleared without residual inflammation; (2) the viral infection persists; or (3) the viral infection leads to autoimmune-mediated inflammation that persists despite viral disappearing. In (2) or (3), patients may progress to chronic dilated cardiomyopathy.^9^ We have already reported that patients with reduced LVEF tend to improve in follow-up.^7^ This study’s multivariate analysis also shows that reduced LVEF was an independent prognostic factor. This suggests that myocardial inflammation may gradually improve with follow-up.

The important finding is that one-third of the FM patients had low PNI at discharge, indicating that patients with persistent inflammation were likely to have a poor prognosis. Additionally, even when including the Barthel index, indicating daily living activity, PNI remained an independent prognostic factor in the multivariate analysis. This finding suggests that PNI is a prognostic factor independent of daily living activity. However, PNI on admission has not indicated a prognosis compared to PNI at discharge. This is strongly influenced by the course of treatment, comorbidities, and length of hospital stay, but there was no significant difference in the content of the treatments. This gap may be due to the different roles of lymphocytes, which are components of PNI, in the acute and chronic phases. Lymphocytes, including the activated T cell system, are indicated to be the major pathophysiological mechanism underlying auto-immune myocarditis and autoimmune inflammatory cardiomyopathy.^25,26,27^ However, a decrease in the number of lymphocytes indicates a decline in immune function and an increase in inflammatory response.^28,29^ This knowledge gap may indicate that myocardial injury is triggered by autoimmune inflammation in FM with the acute phase, while lymphocytes are part of the normal immune function in the chronic phase.

PNI is a simple biomarker derived from blood tests. If we can predict patients’ prognosis with reasonable accuracy, understanding the clinical course and disease progression of patients presenting with myocarditis would facilitate resource management and early implementation of certain therapeutic options, including pharmacologic and mechanical circulatory support.

Whether using blood biomarkers or histological evaluation, it is extremely important to confirm whether inflammation persists in patients with FM. This study can stimulate discussion and verification of the residual inflammation, leading to a debate on whether additional treatment is necessary for the chronic phase of FM patients. Recently, neutrophil blockers for myocarditis have also been developed.^30^ In the future, new immunosuppressive therapies may be guided by biomarkers of inflammation in FM patients. It should be emphasized that, in the acute and chronic phases of FM, evaluation of not only cardiac function and biomarkers but also nutritional status leads to predicting mid-to-long-term outcomes in patients with FM.

## Limitations

Since this study was conducted in Japan, patients’ backgrounds and clinical practices may differ in other countries. This cohort contains non-histologically proven FM patients because we included patients with suspected FM. However, some myocarditis cannot be proven histologically in clinical settings, and PNI is useful clinically because it is less invasive. Thus, we used a more clinical data set with FM. The survival bias limited the outcome because the patients with in-hospital death were excluded. The number of patients with giant cell myocarditis was small in our study, and it will be necessary to increase the number of patients for further investigation on the analysis of PNI according to histological subtypes. The follow-up data were from the medical record review, and the follow-up periods varied across patients. Therefore, some outcomes after the follow-up period could be missed if the follow-up terminates prematurely.

## Conclusions

One-third of discharged FM patients with low PNI had a higher risk of mortality in the chronic phase. PNI at discharge was an independent predictor of all-cause death or rehospitalization with cardiovascular disease, not PNI on admission. This study provokes clinical insight into the phenotype of chronic inflammation in FM and optimal follow-up management with low PNI.

## Data Availability

All data is avallable.

https://center6.umin.ac.jp/cgi-open-bin/ctr_e/ctr_view.cgi?recptno=R000045352

## Non-standard Abbreviations and Acronyms

BNP: brain natriuretic peptide
CRP: C-reactive protein
eGFR: estimated glomerular filtration rate
EMB: endomyocardial biopsy
FM: fulminant myocarditis
JRFM: Japanese registry of fulminant myocarditis
LVEF: left ventricular ejection fraction
NYHA: New York Heart Association
PNI: prognostic nutritional index

## Acknowledgments

We would like to thank all the facilities with the Japanese Registry of Fulminant Myocarditis.

## References

1. Global Burden of Disease Study 2013 Collaborators. Global, regional, and national incidence, prevalence, and years lived with disability for 301 acute and chronic diseases and injuries in 188 countries, 1990-2013: a systematic analysis for the Global Burden of Disease Study 2013. Lancet. 2015;386:743–800. doi:10.1016/S0140-6736(15)60692-4.

2. Kanaoka K, Onoue K, Terasaki S, Nakano T, Nakai M, Sumita Y, Hatakeyama K, Terasaki F, Kawakami R, Iwanaga Y et al. Features and Outcomes of Histologically Proven Myocarditis With Fulminant Presentation. Circulation. 202;146:1425–1433. doi:10.1161/CIRCULATIONAHA.121.058869.

3. Ammirati E, Cipriani M, Moro C, Raineri C, Pini D, Sormani P, Mantovani R, Varrenti M, Pedrotti P, Conca C, et al. Clinical Presentation and Outcome in a Contemporary Cohort of Patients With Acute Myocarditis: Multicenter Lombardy Registry. Circulation. 2018;138:1088–1099. doi:10.1161/CIRCULATIONAHA.118.035319.

4. Ammirati E, Veronese G, Brambatti M, Merlo M, Cipriani M, Potena L, Sormani P, Aoki T, Sugimura K, Sawamura A, et al. Fulminant Versus Acute Nonfulminant Myocarditis in Patients With Left Ventricular Systolic Dysfunction. J Am Coll Cardiol. 2019;74:299–311. doi:10.1016/j.jacc.2019.04.063.

5. Anzini M, Merlo M, Sabbadini G, Barbati G, Finocchiaro G, Pinamonti B, Salvi A, Perkan A, Di Lenarda A, Bussani R, et al. Long-term evolution and prognostic stratification of biopsy-proven active myocarditis. Circulation. 2013;128:2384–2394. doi:10.1161/CIRCULATIONAHA.113.003092.

6. Magnani JW, Dec GW. Myocarditis: current trends in diagnosis and treatment. Circulation. 2006 Feb 14;113(6):876–890. doi:10.1161/CIRCULATIONAHA.105.584532.

7. Kanaoka K, Onoue K, Terasaki S, Nakai M, Iwanaga Y, Miyamoto Y, Saito Y. Changes in Cardiac Function Following Fulminant Myocarditis. Circ Heart Fail. 2024;17:e010840. doi:10.1161/CIRCHEARTFAILURE.123.010840.

8. Liu PP, Mason JW. Advances in the understanding of myocarditis. Circulation. 2001;104:1076–1082. doi:10.1161/hc3401.095198.

9. Tschöpe C, Ammirati E, Bozkurt B, Caforio ALP, Cooper LT, Felix SB, Hare JM, Heidecker B, Heymans S, Hübner N, et al. Myocarditis and inflammatory cardiomyopathy: current evidence and future directions. Nat Rev Cardiol. 202;18:169–193. doi:10.1038/s41569-020-00435-x.

10. Hang W, Chen C, Seubert JM, Wang DW. Fulminant myocarditis: a comprehensive review from etiology to treatments and outcomes. Signal Transduct Target Ther. 2020;5:287. doi:10.1038/s41392-020-00360-y.

11. Ma S, Zhang B, Lu T, Li D, Li T, Shen Z, He C, Wang Y, Li B, Zhang H, et al. Value of the prognostic nutritional index (PNI) in patients with newly diagnosed, CD5-positive diffuse large B-cell lymphoma: A multicenter retrospective study of the Huaihai Lymphoma Working Group. Cancer. 2022;128:3487–3494. doi:10.1002/cncr.34405. Epub 2022 Aug 6.

12. Ding P, Guo H, Sun C, Yang P, Kim NH, Tian Y, Liu Y, Liu P, Li Y, Zhao Q. Combined systemic immune-inflammatory index (SII) and prognostic nutritional index (PNI) predicts chemotherapy response and prognosis in locally advanced gastric cancer patients receiving neoadjuvant chemotherapy with PD-1 antibody sintilimab and XELOX: a prospective study. BMC Gastroenterol. 2022;22:121. doi:10.1186/s12876-022-02199-9.

13. Mullen JL, Buzby GP, Matthews DC, Smale BF, Rosato EF. Reduction of operative morbidity and mortality by combined preoperative and postoperative nutritional support. Ann Surg. 1980;192:604–613. doi:10.1097/00000658-198019250-00004.

14. Wei W, Wu X, Jin C, Mu T, Gu G, Min M, Mu S, Han Y. Predictive Significance of the Prognostic Nutritional Index (PNI) in Patients with Severe COVID-19. J Immunol Res. 2021;2021:9917302. doi:10.1155/2021/9917302.

15. Cheng YL, Sung SH, Cheng HM, Hsu PF, Guo CY, Yu WC, Chen CH. Prognostic Nutritional Index and the Risk of Mortality in Patients With Acute Heart Failure. J Am Heart Assoc. 201;6:e004876. doi:10.1161/JAHA.116.004876.

16. Huang Y, Zhang Q, Li P, Chen M, Wang R, Hu J, Chi J, Cai H, Wu N, Xu L. The prognostic nutritional index predicts all-cause mortality in critically ill patients with acute myocardial infarction. BMC Cardiovasc Disord. 2023;23:339. doi:10.1186/s12872-023-03350-4.

17. Chen QJ, Qu HJ, Li DZ, Li XM, Zhu JJ, Xiang Y, Li L, Ma YT, Yang YN. Prognostic nutritional index predicts clinical outcome in patients with acute ST-segment elevation myocardial infarction undergoing primary percutaneous coronary intervention. Sci Rep. 2017;7:3285. doi:10.1038/s41598-017-03364-x.

18. Caforio AL, Pankuweit S, Arbustini E, Basso C, Gimeno-Blanes J, Felix SB, Fu M, Heliö T, Heymans S, Jahns R, et al. Current state of knowledge on aetiology, diagnosis, management, and therapy of myocarditis: a position statement of the European Society of Cardiology Working Group on Myocardial and Pericardial Diseases. Eur Heart J. 2013;34:2636–2648, 2648a-2648d. doi:10.1093/eurheartj/eht210.

19. JCS Joint Working Group. Guidelines for diagnosis and treatment of myocarditis (JCS 2009): digest version. Circ J. 2011;75:734–743. doi:10.1253/circj.cj-88-0008.

20. Abe A, Hayashi H, Ishihama T, Furuta H. Prognostic impact of the prognostic nutritional index in cases of resected oral squamous cell carcinoma: a retrospective study. BMC Oral Health. 2021;21:40. doi:10.1186/s12903-021-01394-6.

21. Mohri Y, Inoue Y, Tanaka K, Hiro J, Uchida K, Kusunoki M. Prognostic nutritional index predicts postoperative outcome in colorectal cancer. World J Surg. 2013;37:2688–2692. doi:10.1007/s00268-013-2156-9.

22. Hu JR, Florido R, Lipson EJ, Naidoo J, Ardehali R, Tocchetti CG, Lyon AR, Padera RF, Johnson DB, Moslehi J. Cardiovascular toxicities associated with immune checkpoint inhibitors. Cardiovasc Res. 2019;115:854–868. doi:10.1093/cvr/cvz026.

23. Trachtenberg BH, Hare JM. Inflammatory Cardiomyopathic Syndromes. Circ Res. 2017;121:803–818. doi:10.1161/CIRCRESAHA.117.310221.

24. Nagai T, Inomata T, Kohno T, Sato T, Tada A, Kubo T, Nakamura K, Oyama-Manabe N, Ikeda Y, Fujino T, et al. JCS 2023 Guideline on the Diagnosis and Treatment of Myocarditis. Circ J. 202;87:674–754. doi:10.1253/circj.CJ-22-0696.

25. Ohta-Ogo K, Sugano Y, Ogata S, Nakayama T, Komori T, Eguchi K, Dohi K, Yokokawa T, Kanamori H, Nishimura S, et al. Myocardial T-Lymphocytes as a Prognostic Risk-Stratifying Marker of Dilated Cardiomyopathy - Results of the Multicenter Registry to Investigate Inflammatory Cell Infiltration in Dilated Cardiomyopathy in Tissues of Endomyocardial Biopsy (INDICATE Study). Circ J. 2022;86:1092–1101. doi:10.1253/circj.CJ-21-0529.

26. Anzai A, Mindur JE, Halle L, Sano S, Choi JL, He S, McAlpine CS, Chan CT, Kahles F, Valet C, et al. Self-reactive CD4+ IL-3+ T cells amplify autoimmune inflammation in myocarditis by inciting monocyte chemotaxis. J Exp Med. 2019;216:369–383. doi:10.1084/jem.20180722.

27. Opavsky MA, Penninger J, Aitken K, Wen WH, Dawood F, Mak T, Liu P. Susceptibility to myocarditis is dependent on the response of alphabeta T lymphocytes to coxsackieviral infection. Circ Res. 1999;85:551–558. doi:10.1161/01.res.85.6.551.

28. Verdon DJ, Mulazzani M, Jenkins MR. Cellular and Molecular Mechanisms of CD8+ T Cell Differentiation, Dysfunction and Exhaustion. Int J Mol Sci. 2020;21:7357. doi: 10.3390/ijms21197357.

29. Wan YY. Multi-tasking of helper T cells. Immunology. 2010;130:166–171. doi: 10.1111/j.1365-2567.2010.03289.x.

30. Carai P, González LF, Van Bruggen S, Spalart V, De Giorgio D, Geuens N, Martinod K, Jones EAV, Heymans S. Neutrophil inhibition improves acute inflammation in a murine model of viral myocarditis. Cardiovasc Res. 2023;118:3331–3345. doi: 10.1093/cvr/cvac052.

